# Exploring the impact of integrated health and social care services on child health and wellbeing in underserved populations: a systematic review

**DOI:** 10.1101/2024.01.05.24300706

**Authors:** Chris Bird, Lorraine Harper, Syed Muslim, Derick Yates, Ian Litchfield

## Abstract

**Objective:** To explore the evidence for interventions that integrate child health and social care and support programmes and the impact they have on child health and wellbeing.

**Data sources:** The Cochrane Library, Ovid Medline, Ovid Embase, Ovid Emcare, Ovid Health Management Information Consortium (HMIC) database, and Ovid Social Policy and Practice, Proquest Psychinfo and Ebscohost Cinahl.

**Eligibility:** Peer-reviewed original research that described an intervention integrating health care and social support or care interventions for children and young people (CYP) up to the age of 18 years in high-income countries. All databases were searched from inception to August 2023.

**Data extraction and synthesis:** 16 studies were identified: 4 RCTs, 5 quantitative studies, 5 qualitative studies and 2 mixed methods studies. A narrative review and quality check of included studies was performed. Study heterogeneity meant a meta-analysis could not be completed.

**Results:** Five qualitative, five quantitative, two mixed methods and four randomised controlled trials were included. We identified three main models of delivering integrated health and social care services: targeted support for vulnerable groups, where the provision of packages of interventions focussed on target populations showed potential for decreasing the need for social support in the long-term but with limited evidence for reducing referrals into other services. They were more successful in meeting specific objectives such as lower rates of smoking, and reducing repeat pregnancies; collaborative health and social support were typically collocated services which improved collaborative working but with little impact on workload, job satisfaction, or service delivery; and school centred health and social care, which improved some aspects of CYP wellbeing and physical health but with concerns they added to teacher workload.

**Conclusions:** Integrated health and social support programmes offer promising solutions to addressing health inequity in children and young people in underserved populations. However, more robust and consistent study designs are needed to guide researchers and policy makers in their implementation and evaluation.

What is already known on this topic.

- Integrated care that is equipped to mitigate at least some of the social determinants of health is considered key to improving health inequalities in children and young people in underserved populations.
- Despite increasing investment in integrating health and social support there is little evidence of which models of delivery are most effective in which circumstances nor of the precise impact on target groups and the wider healthcare system.

What this study adds

- We identified three models of integrating health and social support: Packages of interventions targeted at specific groups; Collaborative (and collocated) health and social support; and School-centered health and social care interventions.
- Results across the three models offered promise of improved care and support for the underserved, referral rates into other services tended to be lower and packages interventions achieved positive results for specific objectives. However, findings were inconsistent, drop-out rates were high, and there are concerns over sustainability without sufficient resources.

How this study might affect research, practice and policy

- Embedding iterative co-production in future research and interventions could improve engagement and outcomes and it’s important that further works explores their cost effectiveness and implications for other elements of health and care services.
- Appropriate resources and a longer-term commitment to promoting integrated health and social support is needed to fully understand the strengths and weaknesses of the offer and maximise the potential benefits.

## Introduction

Children, young people (CYP) and their families living in high income countries face mounting challenges to their health and well-being, as the prevalence of chronic conditions, obesity, and mental ill health continues to increase (1). These challenges are exacerbated in underserved populations i.e., minoritized and economically-deprived communities (2, 3), by a range of socio-economic and cultural pressures that inhibit access and utilisation of primary or preventative health care services (4–7). This has led to a widespread rise in children’s attendances to emergency departments frequently due to conditions that could be more effectively treated in community settings (8–12).

The need for more responsive, culturally sensitive care for CYP from underserved populations has led to efforts in North America, Europe and Australia to prioritise more localized service delivery that integrates several strands of health and social care and places a greater emphasis on public and preventative health (13–19). The integrated services that have emerged are delivered by various combinations of health care providers, social care practitioners, community advocates, and public institutions, and situated in a range of central and localised clinical and locality-based settings (20, 21). Together they share the aim of providing widely accessible health and social care for CYP and their families that can help treat and manage acute and chronic health care alongside the necessary social support that can help mitigate the social determinants of ill-health such as poor housing, domestic violence, or food poverty (21, 22).

However, despite widespread investment in these systems in countries such as the United Kingdom (23), evidence of the benefits of integrating health and social care remains inconsistent, particularly amongst underserved CYP (24, 25): Little is known of which integrated models are most effective, including the precise combination of services, the specific outcomes they improve, or the impact on the surrounding health economy (26, 27). This systematic review is the first that has collated and examined the impact of these integrated health and social care services on CYP in underserved populations. It suggests a typography of the various service models employed and presents the qualitative and quantitative evidence of the effectiveness of each.

## Methods

### Study design

This work consists of a mixed methods systematic review of qualitative and quantitative studies (28). We used the PerSPEcTiF model to frame the review question (see Table 1) (29) and followed the Preferred Reporting Items for Systematic Reviews and Meta-Analyses (PRISMA) guideline (30). The study is registered on PROSPERO (International Prospective Register of Systematic Reviews: CRD42023399907) (31).

**Table 1:** Review framework using PerSPEcTiF (*Booth et al, 2019*)

| Full Review Question (using PerSPeCTiF framework) |  |
| --- | --- |
| Do interventions that integrate child health and social support intervention impact child health and wellbeing? |  |
| Perspective | From the perspectives of both families and CYP who use the service, those who deliver the service and outside observers. |
| Setting | Interventions where healthcare and social support programmes for children and young people (CYP) are integrated, including qualitative studies, quantitative studies (RCTs, cohort, observational, quasi experimental), no date limit (exclude case reports, reviews, commentary). |
| Phenomenon of interest/problem | Impacts on a wide range of outcomes on child health (preventive, acute, chronic health issues) and wellbeing (eg school attendance, domestic violence). |
| Environment | High income countries with a particular focus on interventions in deprived areas, to focus on settings similar to the UK (eg Europe, New Zealand, Australia). |
| **Comparison** | Standard care, if a comparator available. |
| **Time** | Children and young people (CYP) <18 years, at any point during their childhood (eg infant, pre-school, primary and secondary school age). |
| **Findings** | Impacts on child health and wellbeing, eg school attendance, asthma control, including: qualitative – patient/professional value/experience of service; quantitative – cost effectiveness, primary and secondary care use, school attendance. Given the complex nature of these interventions, outcome measures likely to be heterogeneous. |

### Search strategy

The review question was designed using the PerSPEcTiF question framework, to enable the search to best identify a set of relevant abstracts of interest, and the database search structure followed a Population, Exposure, Outcomes (PEO) approach (see Supplementary File 1). The following databases were searched: Cochrane Library, Ovid Medline, Ovid Embase, Ovid Emcare, Ovid Health Management Information Consortium (HMIC) database, Ovid Social Policy and Practice, Proquest Psychinfo and Ebscohost Cinahl.

### Inclusion criteria

Studies were eligible for inclusion if their focus was integrated health and social care and support approaches, in “underserved populations” defined as those groups possessing “health differences that are avoidable, unnecessary, and unjust” (32). All databases were searched from inception to August 2023 with no limits in relation to study, publication type, language or date of publication were applied. The search identified a combination of relevant subject headings within those databases using a controlled vocabulary; MeSH in Cochrane, Medline and Cinahl. Emtree in Embase and Emcare and APA Thesaurus of Psychological Index Terms in PsychInfo combined with keywords and free text word variations. Proximity operators were used to maximise the efficiency of the search strategy when searching for phrase variations. The full search strategy is available in Supplementary File 1.

### Study selection and assessment of quality and bias

Identified studies were collated and managed using Endnote and Covidence software (33, 34). Two independent reviewers (CB and SM) identified relevant papers by reading titles and abstracts and disagreements were resolved through joint review and consensus. Both reviewers assessed study quality and risk of bias, using the Critical Appraisal Skills Programme checklist for qualitative studies (35), the Effective Public Healthcare Panacea Project’s quality assessment tool for quantitative studies (36) and McGill University’s Mixed Methods Appraisal Tool for mixed methods studies(37).

## Data Analysis

Data were collated, organised, and analysed according to the shared characteristics of the service they delivered. If the data was a available a meta-analysis of patient outcomes would be conducted, in its absence a narrative synthesis using qualitative data augmented with quantitative data where available (29, 37).

## Results

### Study characteristics

A total of 3,741 studies were imported for screening and four studies were found via hand searches. 1,421 duplicates were removed, 3,701 studies were screened, 43 full text studies were assessed for eligibility and 16 studies were included in the review. Studies were excluded because they were either of an incorrect intervention (n=13), study design (n=4), setting (n=1), outcome (n=1), population (n=1), or reported no results of impact (n=7). These are described in the PRISMA flow diagram (see Figure 1) (30). Five qualitative, five quantitative, two mixed methods and four randomised controlled trials were included. All studies were carried out in Australia, North America, or Western Europe. The characteristics of each study are further described in Table 2.

**Figure 1:**
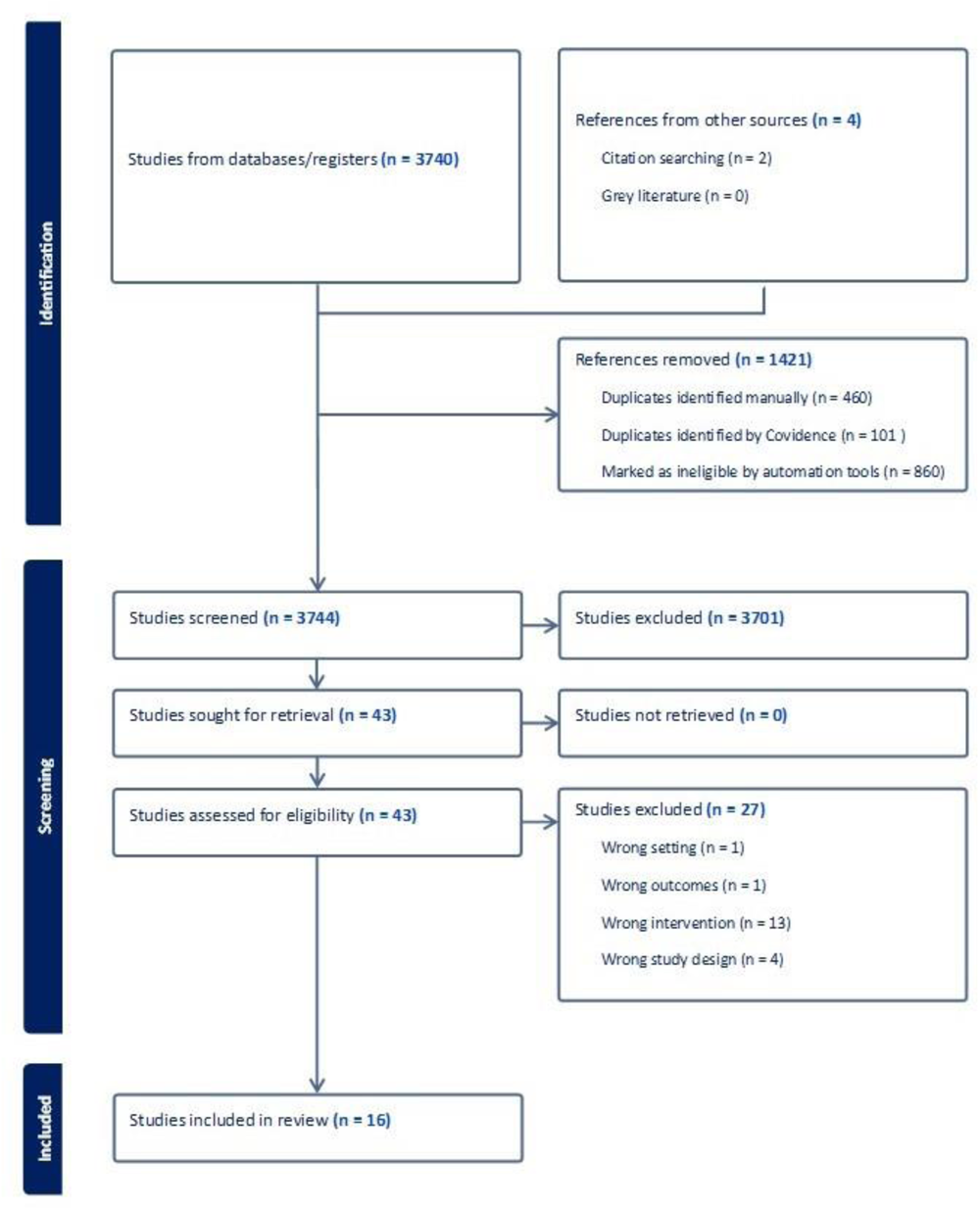
PRISMA flow diagram.

**Table 2:**
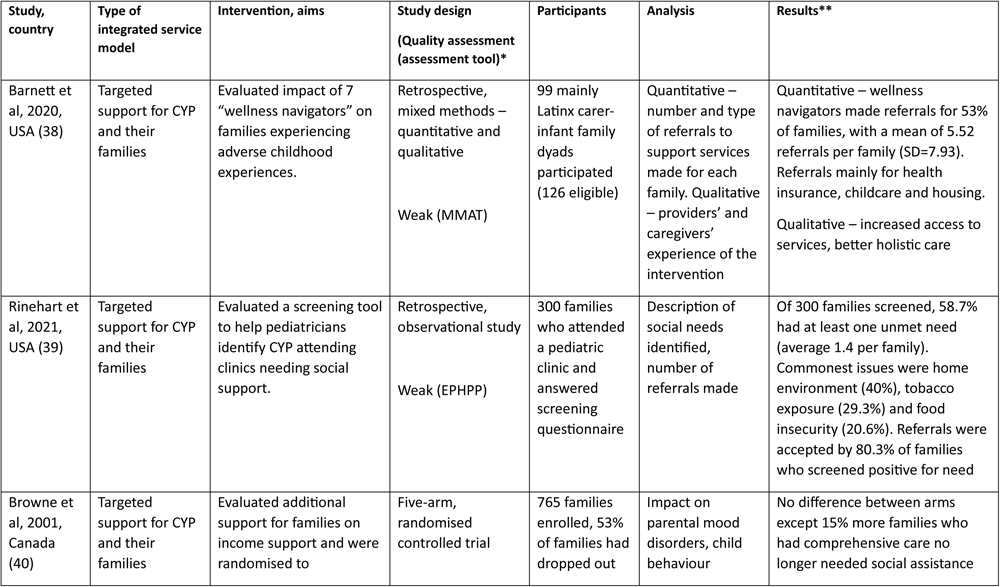
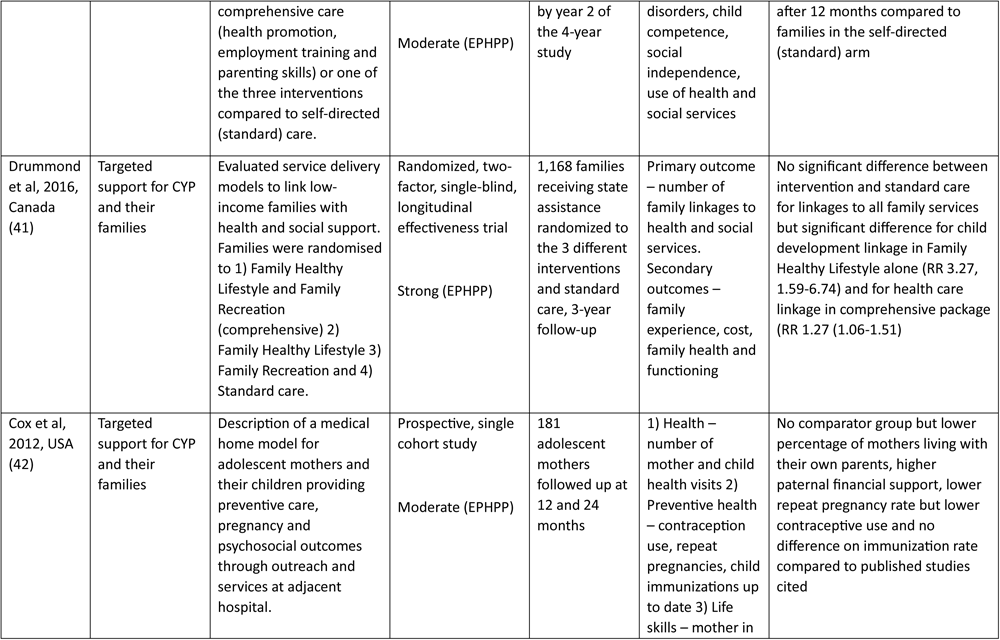
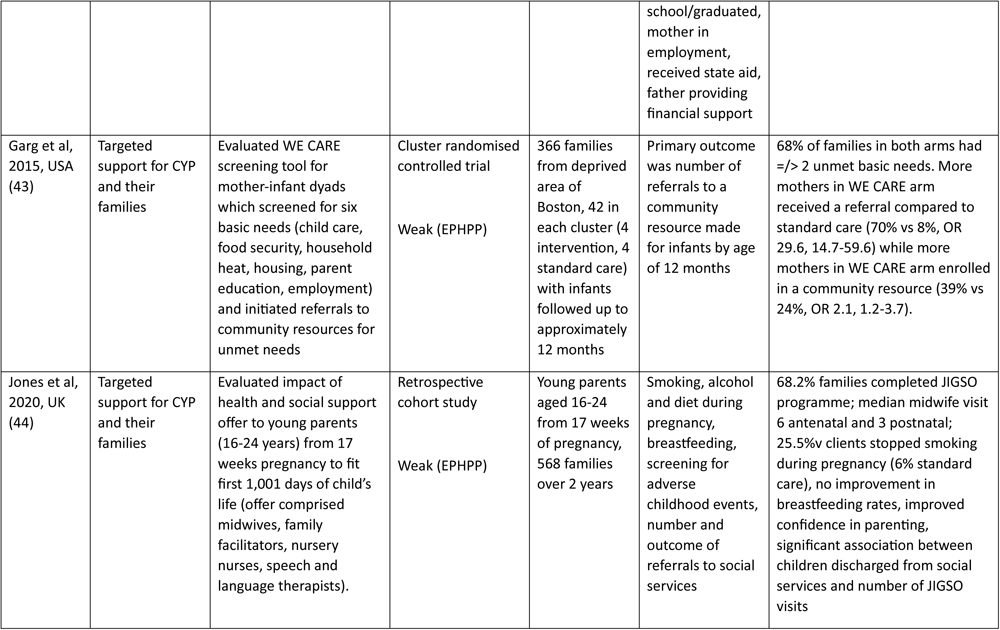
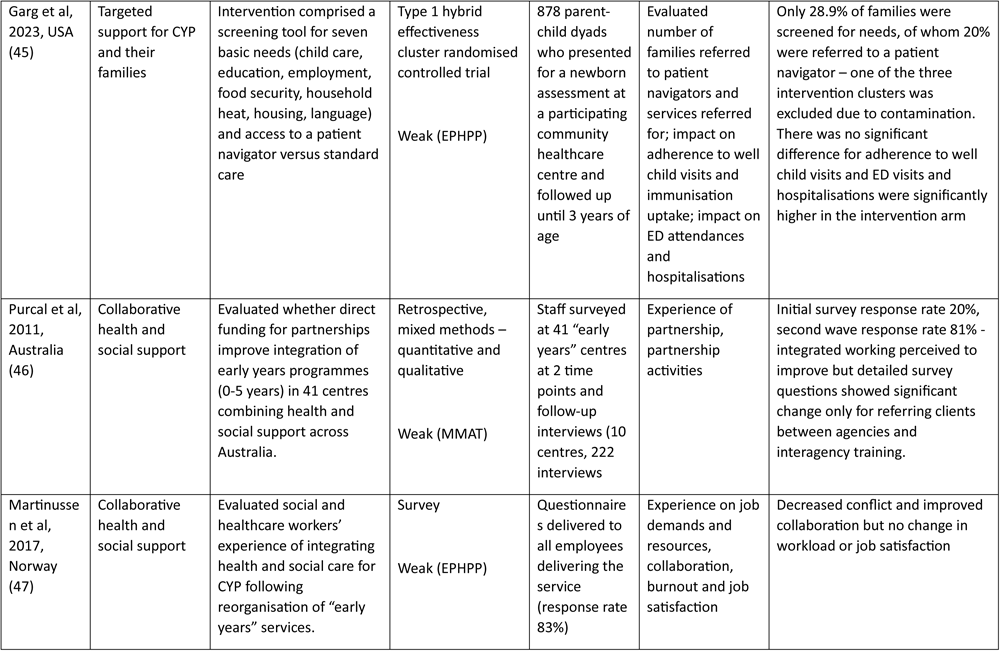
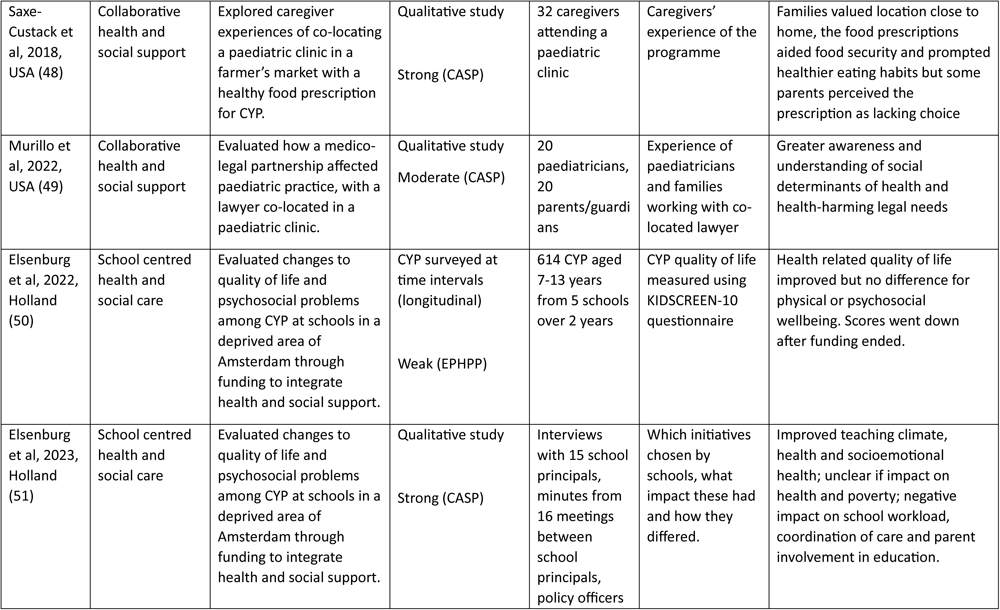
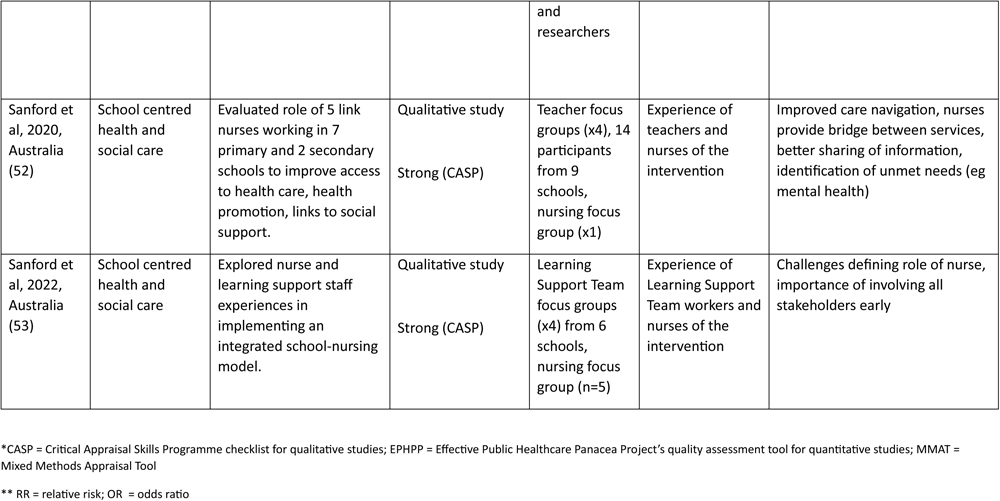
Characteristics of included studies.

### Integrated health and social care models

We identified three types of integrated health and social care services: *Targeted support for CYP and their families*, where specified groups with additional needs were identified and then receive a range of health and care interventions (38–45); *Collaborative health and social support*, involving health and social care organisations work together to deliver shared and complementary services (46–49); and *School centred health and social care*, in which health and social care services embedded or directly linked to educational facilities (50–53). A summary of the key characteristics of each of the types of integrated care model can be found in Table 3 and below we summarise the results of the studies identified within each.

**Table 3:** Summary of the models of integrated health and social care designed to support underserved CYP.

| **Integrated model** | **Definition** | **Location** | **Key characteristics** | **Content of intervention(s)** |
| --- | --- | --- | --- | --- |
| ***Targeted support for CYP and families*** | A service based that identified a particular target group or population and then offered one, or a combination of several interventions that combined health and social care intended to impact a pre-specified outcome | Clinical, community, or domestic settings | Identification of those with direct responsibility for CYP health i.e., individual CYP, their carers, and/or families.<br>Single, or a package of, interventions delivered by multiple care organisations reflective of clinical and social need of target groups | Individual packages include numeracy and literacy education, employment training, parenting skills, health promotion, smoking cessation, vaccination, contraception |
| ***Collaborative health and social support*** | An integrated service provided by health and social care organisations and their practitioners offered to localised populations | Predominantly collocated in a shared (community) health care facility | The sharing of key aims, infrastructure, and financial responsibilities across health and social care organisations | A wide range of (preventative) health and social care and support. Including clinical care, legal counselling, lifestyle advice, oral hygiene, and mental health services |
| ***School centred health and social care*** | Health and social care services embedded or otherwise linked with the delivery of primary and secondary education to populations that include CYP from underserved groups | Predominantly delivered within primary and secondary school premises | Health and social care practitioners and/or public health initiatives embedded within or linked to local schools | Programmes that connect CYP and their parents with social and culturally sensitive (preventative) health care and support. Including advice on healthy lifestyle behaviours, support for well-being and signposting to health and social care services |

### Targeted support for CYP and their families

Target groups consisted of CYP and their families (39–41), or (young) mothers with infants (38, 42–45). They were identified via bespoke screening tools (43, 45), through their existing or previous use of social care or support (38), and an actual or proxy measure of low-income or deprivation (39–41). The interventions were typically delivered in community based care centres or clinics (38, 39, 43–45), or in two instances the CYP’s home (42, 44). Four studies targeted CYP and their families, three were recognised as requiring additional needs by direct or proxy measure of deprivation (39–41) and one by previous contact with social care services (38).

At a single clinic in a deprived district in East Harlem just under 60% of participants had at least one unmet need relating to housing, tobacco exposure, or food insecurity with 80% successfully referred to the appropriate social support as a result (39). A multi-arm randomised controlled trial (RCT) set in Canada identified vulnerable families by a locality-based deprivation score that accessed a range of interventions including various combinations of health promotion, parenting skills, and employment training, with the published interim analysis indicating that those receiving the intervention were less likely to need social assistance 12 months later (40). A second RCT, also in Canada, recruited participants from locality-based deprivation scores described the impact of a range of family-based lifestyle and recreational interventions with significant improvements in engagement with child development services (RR 3.27, 1.59-6.74) and health care (RR 1.27 (1.06-1.51) (41). However, over half of families receiving the intervention dropped out after two years and the authors observed that integrating the work of the existing agencies did not address longstanding shortages in service capacity (41). Barnett et al reported that Latinx carer-infant dyads identified by previous contact with social support subsequently had increased referrals to organisations providing health insurance, childcare and housing (38).

Two studies targeted young mothers (42, 44): One, set in the USA, that integrated support from hospital staff and social workers into a “medical home” model and reported they were less likely to live with their own parents, have a repeat pregnancy and received greater paternal financial support (42). The other study set in Wales (UK) consisted of health and social support from a team of midwives, family facilitators, nursery nurses, and speech and language therapists (44). They reported reduced smoking rates, and improved confidence in parenting though no increase in breastfeeding (44).

Two related studies targeted mothers (of any age) and their infants with unmet needs (43, 45). They found that those identified using the tool were significantly more likely to receive a referral to community (social) services, though only half of those actually received additional support (43). Adapting the screening tool to incorporate multiple languages and linking participants with a patient-peer navigator increased the likelihood of an ED visit or hospitalisation (45).

### Collaborative health and social support

The facilitated collaborations consisted of community-based co-located social support and health care services set in Australia (46), USA (48, 49), and in Norway an organisational-level collaborative service (47). Purcal et al’s Australian study of state funding for integrating professional health and social support found a significant increase in inter-agency referrals but no impact on planning, service delivery or co-location, according to senior managers, managers and frontline staff (46).

Two single-centred, US-based studies described social support interventions co-located in paediatric clinics: one provided a fresh food prescription though families felt that they aided food security but also felt the options were constrictive and would have preferred vouchers (48). In the second a paediatric clinic provided a lawyer to tackle health-harming legal needs such as those relating to housing, utilities, guardianship, education and benefits (49). The qualitative data indicated greater confidence and trust, from CYP and families, in clinical staff, who in turn reported improved awareness and understanding of the social determinants of their patients’ health (49). Martinussen et al’s survey of Norwegian health professionals following the re-organisation of services to better integrate social care found improved collaboration but did little to improve job satisfaction or reduce workload (47).

### School centred health and social care

Of the four studies identified, two evaluated a single intervention in Australia that comprised of linked nurses within primary and secondary schools situated within economically disadvantaged locations in Australia evaluating (52, 53); and two in the Netherlands exploring the impact of central funding on a range of small scale health and social care packages determined by primary school leaders (50, 51).

The Australian qualitative studies explored the views of nursing link workers and teachers and learning support workers working together in primary and secondary schools in Australia (52, 53). Both school and nursing staff reported that care navigation improved, with better information sharing and identification of unmet needs (eg mental health) but there were challenges in defining the nurses’ roles and how they worked alongside school support staff (52, 53).

The two studies that evaluated the 2-year programme in the Netherlands, where government provided €125,000 to schools in economically disadvantaged areas to fund their choice of interventions with the premise they would integrate health and social support for CYP aged 7-13 years (50, 51). The first described the results of a longitudinal survey of pupils which indicated that health related quality of life and psychosocial problems improved, and though the scores used to measure wellbeing displayed little variation over time, they did drop off once funding ended (50). The second study of stakeholder perceptions, described perceptions of improved wellbeing, physical health and classroom behaviour, though the school leaders were concerned about sustained impact due to the impact of interventions on teacher workload, and coordination of care (51).

## Discussion

### General findings

This review provides valuable and novel insight into the various attempts at integrating health and social care for the benefit of CYP and families from underserved populations. Of the papers identified, various integrated service offers emerged, and we were able to produce a typology that categorised the studies we identified into three different models of service delivery. *Targeted support for CYP and their families:* Which first involved identification of vulnerable groups and then the subsequent provision of packages of interventions targeted at their specific needs showed potential for decreasing the need for social support in the long-term, but with limited evidence for improving intervention specific outcomes such as referrals into other services, lower rates of smoking, reduced repeat pregnancies and greater likelihood of discharge from social services. *Collaborative health and social care* found some improvement in collaboration between previously disparate services, but no impact on workload, job satisfaction or service delivery. There was limited evidence of the benefits of co-locating social support in paediatric clinics, and collocated legal assistance offered promise.

*School centred health and social care* improved some aspects of CYP wellbeing and physical health, but senior educational staff reported that additional funds failed to improve school and social care support and increased teacher workload.

## Specific findings

### Targeted support for CYP and families

Target groups were readily identified but the evidence of the various approaches used to identify these groups was inconsistent, with some improvements reported in streamlining referrals into other services (39, 40, 43, 45), reducing the number of repeat pregnancies (44), or smoking (48). Similar approaches targeting deprived families (though without integrating health and social care), have also shown promise in promoting healthy behaviours for example raising awareness of oral health and reduced obesity (20, 54–56).

The lack of more definitive evidence of such health and social care interventions designed to impact the underserved might be attributed in part to broader systemic difficulties in engaging with these groups due to their frequent changes of address; lack of trust in centralised authorities, and language and cultural barriers (57). To overcome some of these pervasive socio-cultural barriers to accessing health and social care there is a need to recognise the value of using alternative means of improving the outreach to the most vulnerable such as housing associations (58), (59) or homelessness charities (60). Having done so, co-design initiatives of health and social care interventions can become more meaningful and genuinely increase ownership of the intervention whilst reducing the stigma of accessing support (61). Future interventions might also be better supported by embedding peers or community connectors in the delivery or facilitation of the service, to help address the persistent issues of mistrust and engagement with mainstream health and social support services (62, 63).

### Collaborative health and social support

The studies we uncovered reported limited benefits of collaborative health and social care including more effective referral into social support services and increased job satisfaction (46–49). The majority of previous work that has explored inter agency collaborative working has focused on creating teams of primary and secondary care clinicians (64, 65). What remains less well understood is how best to combine health and social care services (24, 25).

The colocation of existing services is widely recognised as a successful way of integrating multi-disciplinary health care teams: improving communication, understanding and mutual learning (66, 67). However, professional partnership working requires bridging differences in training, aims, and work practices of health and social care practitioners (68). If the integration of health and social care services is to be sustained in the long-term then fundamental issues around professional identities and boundaries need to be addressed (69). This requires changes in training and education to better ensure such partnership working remains safe and effective (70) with techniques such as inter-disciplinary observation recognised as an aid to fostering mutual respect, greater job satisfaction and workforce retention (71–74).

### School centred health and social care

Delivery of health and wellbeing through schools has been promoted globally for several decades and was recognised as part of the World Health Organisation’s 1986 Charter on Health Promotion which asks that schools constantly strengthen their capacity as a “healthy setting for living, learning and working” (75). In the studies we identified, attempts at achieving similar aims involved either integrating health and social care practitioners into the school workforce (52, 53), or by using additional funds to finance a number of health promotion interventions around diet and exercise (50, 51). That both approaches reported positive effects on health and well-being but with negative consequences on teacher workload (50–53) reflects the findings of other types of school-based health interventions (76, 77). In these cases they reported promising improvements in anxiety, mental health, asthma management and vision screening (76) but with the impact on educational outcomes and constraints of staff resource hindering sustainability (77). There is also a more fundamental issue that such school-based interventions fail to address, which is that their attempts at reaching underserved populations is predicated on their regular attendance at school, which is usually below national averages (78) but since COVID-19 the number of children in the UK from disadvantaged backgrounds regularly attending school has fallen (79).

### Strengths and limitations

This systematic review was prospectively registered and the identification of studies conducted with reference to best practice by two researchers working independently (30, 80). Despite the comprehensive search strategy identifying 16 papers the overall quality of the evidence was poor, only four of the studies were RCTs (40, 41, 43, 45) and there was little data on outcomes and impact over the medium and long term,. The findings were further limited by high drop-out rates (40–42, 44, 45) and a lack of homogenenity even with model types precluded a formal meta-analysis and any meaningful comparison of the effectiveness of the three model types.

### Implications for policy and research

In light of growing child poverty rates in high income environments (81, 82), the lack of sustained engagement and high-drop-out rates reported by many of our studies (40–42, 44, 45) highlights the importance of delivering services co-designed with intended users (41, 45, 83). The three typologies of integrated service we identified are not intended to be a definitive list and others may emerge including hybrid service offerings that combine elements of each. However, the importance of effective system navigation was understood across all models (40, 42–45, 48), and its importance in accessing and engagement with care is widely recognised both in the UK and elsewhere (9, 84–86).

The establishment of a more robust evidence base is inhibited by the current focus on short-term pilots and funding cycles despite complex interventions needing time to become embedded and medium term outputs that extend beyond the limitations of annual funding cycles (15, 87). Their evaluation also needs to incorporate more precise description of the service model, and the measurement of outcomes valued to both service and patient using mixed methodologies and some element of cost effectiveness (88, 89).

## Conclusion

While we lack robust evidence of the benefits of integrating health and social support for CYP from underserved populations there are promising signs of positive impacts but to fully understand their potential, more robust evaluation methods are needed of services that are commissioned for longer periods of time and retain maximum flexibility in their attempts to engage with underserved communities.

## Author’s statement

## Data Availability

Data are available upon reasonable request. Additional data and materials such as data collection forms, data extraction and analysis templates and can be obtained by contacting the corresponding author.

## Acknowledgements

N/A

## Contributors

## Patient consent

N/A

