## Supplementary file 1 for "Exploring the impact of integrated health and social care services on child health and wellbeing in underserved populations: a systematic review"

***Supplementary File 1 Population, exposure, outcome search strategy + search terms***

**Population, exposure, outcome search strategy**

| Search component | Search strategy |
| --- | --- |
| Population | All papers on children and young people up age 18 years. |
| Exposure | All papers on the topic of integrated healthcare, intersectoral collaboration or community-based health care. We also searched for specific integrated healthcare programmes which had been identified by the review team. |
| Outcomes | Reported outcomes for health and wellbeing, with a focus on health inequalities or inequality in health care, health status or social determinants of health. |

**Search terms**

Ovid MEDLINE(R) ALL <1946 to January 30, 2023>

1	*"Delivery of Health Care, Integrated"/ and exp Cooperative Behavior/	702

2	exp Intersectoral Collaboration/	2568

3	(exp Community Health Services/ or exp Community Mental Health Services/) and exp "Delivery of Health Care, Integrated"/	2256

4	(neighbo?rhood care or neighbo?rhood team*).ab,ti.	23

5	(integrat* adj3 health adj3 (care or service* or team* or group or groups or partners* or organi?ation* or consortium* or collaborat*)).ab,ti.	7898

6	Integrated care partnership.ab,ti.	2

7	Integrated care system.ab,ti.	142

8	Place-based partnerships.ab,ti.	0

9	Provider collaboratives.ab,ti.	0

10	healthcare partnership.ab,ti.	35

11	care partnership.ab,ti.	143

12	community partnership.ab,ti.	714

13	((care or healthcare) adj3 integrated).ab,ti.	15389

14	triple integration.ab,ti.	7

15	"Integrated Care Systems".kw.	16

16	"health and social care partnership".ab,ti.	5

17	"primary and secondary care integration".ab,ti.	6

18	horizontal integration.ab,ti.	201

19	vertical integration.ab,ti.	612

20	shared care.ab,ti.	1526

21	1 or 2 or 3 or 4 or 5 or 6 or 7 or 8 or 9 or 10 or 11 or 12 or 13 or 14 or 15 or 16 or 17 or 18 or 19 or 20	27284

22	exp adolescent/ or exp child/ or exp infant/	3920446

23	(child* or adolescen* or juvenile* or teenage* or youth* or infant* or boy* or girl* or p?ediatric* or young people or young person* or young adult* or youngster* or minor or minors).ab,jn,ti.	2859065

24	"improving the health of children and young people".ab,ti.	2

25	22 or 23 or 24	4881463

26	21 and 25	5370

27	early years.mp.	4699

28	((early or prompt or timely or "in time" or expedient or expeditious or anticipat*) adj3 interven*).mp.	59354

29	((early or prompt or timely or "in time" or expedient or expeditious or anticipat*) adj3 support).mp.	7086

30	((early or earlier) adj3 help*).mp.	8971

31	exp early intervention, educational/ or exp early medical intervention/	6837

32	(head start or sure start or headstart or surestart).ab,ti.	1868

33	("first 1000 days" or "first thousand days" or "1st thousand days" or "1st 1000 days").mp.	429

34	27 or 28 or 29 or 30 or 31 or 32 or 33	80212

35	26 and 34	184

Embase <1974 to 2023 January 30>

1	exp integrated health care system/	13063

2	exp cooperation/	65897

3	exp intersectoral collaboration/	3604

4	(exp community care/ or exp community mental health service/) and exp integrated health care system/	1085

5	(neighbo?rhood care or neighbo?rhood team*).ab,ti.	35

6	(integrat* adj3 health adj3 (care or service* or team* or group or groups or partners* or organi?ation* or consortium* or collaborat*)).ab,ti.	9982

7	Integrated care partnership.ab,ti.	9

8	Integrated care system.ab,ti.	210

9	Place-based partnerships.ab,ti.	0

10	Provider collaboratives.ab,ti.	1

11	healthcare partnership.ab,ti.	39

12	care partnership.ab,ti.	262

13	community partnership.ab,ti.	856

14	((care or healthcare) adj3 integrated).ab,ti.	21843

15	triple integration.ab,ti.	5

16	"Integrated Care Systems".kw.	12

17	"health and social care partnership".ab,ti.	16

18	"primary and secondary care integration".ab,ti.	6

19	horizontal integration.ab,ti.	248

20	vertical integration.ab,ti.	701

21	shared care.ab,ti.	2492

22	1 or 2 or 3 or 4 or 5 or 6 or 7 or 8 or 9 or 10 or 11 or 12 or 13 or 14 or 15 or 16 or 17 or 18 or 19 or 20 or 21	108029

23	exp adolescent/ or exp juvenile/ or exp child/ or exp infant/	3926851

24	(child* or adolescen* or juvenile* or teenage* or youth* or infant* or boy* or girl* or p?ediatric* or young people or young person* or young adult* or youngster* or minor or minors).ab,jn,ti.	3582978

25	"improving the health of children and young people".ab,ti.	2

26	23 or 24 or 25	5158950

27	22 and 26	16921

28	early years.mp.	5892

29	((early or prompt or timely or "in time" or expedient or expeditious or anticipat*) adj3 interven*).mp.	97236

30	((early or prompt or timely or "in time" or expedient or expeditious or anticipat*) adj3 support).mp.	10215

31	((early or earlier) adj3 help*).mp.	14066

32	exp early intervention/	31581

33	(head start or sure start or headstart or surestart).ab,ti.	1995

34	("first 1000 days" or "first thousand days" or "1st thousand days" or "1st 1000 days").mp.	553

35	28 or 29 or 30 or 31 or 32 or 33 or 34	127001

36	27 and 35	514

Ovid Emcare <1995 to 2023 Week 03>

1	exp integrated health care system/	2368

2	exp cooperation/	24860

3	exp intersectoral collaboration/	574

4	(exp community care/ or exp community mental health service/) and exp integrated health care system/	233

5	(neighbo?rhood care or neighbo?rhood team*).ab,ti.	17

6	(integrat* adj3 health adj3 (care or service* or team* or group or groups or partners* or organi?ation* or consortium* or collaborat*)).ab,ti.	5376

7	Integrated care partnership.ab,ti.	1

8	Integrated care system.ab,ti.	105

9	Place-based partnerships.ab,ti.	0

10	Provider collaboratives.ab,ti.	1

11	healthcare partnership.ab,ti.	19

12	care partnership.ab,ti.	114

13	community partnership.ab,ti.	589

14	((care or healthcare) adj3 integrated).ab,ti.	10554

15	triple integration.ab,ti.	1

16	"Integrated Care Systems".kw.	12

17	"health and social care partnership".ab,ti.	11

18	"primary and secondary care integration".ab,ti.	3

19	horizontal integration.ab,ti.	116

20	vertical integration.ab,ti.	288

21	shared care.ab,ti.	1087

22	1 or 2 or 3 or 4 or 5 or 6 or 7 or 8 or 9 or 10 or 11 or 12 or 13 or 14 or 15 or 16 or 17 or 18 or 19 or 20 or 21	42011

23	exp adolescent/ or exp juvenile/ or exp child/ or exp infant/	779682

24	(child* or adolescen* or juvenile* or teenage* or youth* or infant* or boy* or girl* or p?ediatric* or young people or young person* or young adult* or youngster* or minor or minors).ab,jn,ti.	1067677

25	"improving the health of children and young people".ab,ti.	2

26	23 or 24 or 25	1248037

27	22 and 26	5997

28	early years.mp.	2446

29	((early or prompt or timely or "in time" or expedient or expeditious or anticipat*) adj3 interven*).mp.	32374

30	((early or prompt or timely or "in time" or expedient or expeditious or anticipat*) adj3 support).mp.	3798

31	((early or earlier) adj3 help*).mp.	3390

32	exp early intervention/	11628

33	(head start or sure start or headstart or surestart).ab,ti.	1624

34	("first 1000 days" or "first thousand days" or "1st thousand days" or "1st 1000 days").mp.	287

35	28 or 29 or 30 or 31 or 32 or 33 or 34	42704

36	27 and 35	193

HMIC Health Management Information Consortium <1979 to November 2022>

1	exp Integrated care/	2793

2	exp Collaboration/	6857

3	exp Mental health services/ and (exp Integrated care/ or exp Collaboration/)	590

4	(neighbo?rhood care or neighbo?rhood team*).ab,ti.	40

5	(integrat* adj3 health adj3 (care or service* or team* or group or groups or partners* or organi?ation* or consortium* or collaborat*)).ab,ti.	1069

6	Integrated care partnership.ab,ti.	4

7	Integrated care system.ab,ti.	49

8	Place-based partnerships.ab,ti.	5

9	Provider collaboratives.ab,ti.	7

10	healthcare partnership.ab,ti.	12

11	care partnership.ab,ti.	71

12	community partnership.ab,ti.	24

13	((care or healthcare) adj3 integrated).ab,ti.	2049

14	triple integration.ab,ti.	3

15	"health and social care partnership".ab,ti.	26

16	"primary and secondary care integration".ab,ti.	4

17	horizontal integration.ab,ti.	20

18	vertical integration.ab,ti.	41

19	shared care.ab,ti.	340

20	1 or 2 or 3 or 4 or 5 or 6 or 7 or 8 or 9 or 10 or 11 or 12 or 13 or 14 or 15 or 16 or 17 or 18 or 19	11003

21	exp Young people/ or exp children/ or exp adopted children/ or exp boys/ or exp children born early in marriage/ or exp children in care/ or exp eldest child/ or exp girls/ or exp grandchildren/ or exp infants/ or exp older children/ or exp orphans/ or exp pre school children/ or exp schoolchildren/ or exp stepchildren/ or exp unwanted children/ or exp youngest child/	27673

22	(child* or adolescen* or juvenile* or teenage* or youth* or infant* or boy* or girl* or p?ediatric* or young people or young person* or young adult* or youngster* or minor or minors).ab,jn,ti.	43001

23	"improving the health of children and young people".ab,ti.	3

24	21 or 22 or 23	47776

25	exp Preventive measures/ or exp Sure start/	27900

26	early years.mp.	352

27	((early or prompt or timely or "in time" or expedient or expeditious or anticipat*) adj3 interven*).mp.	871

28	((early or prompt or timely or "in time" or expedient or expeditious or anticipat*) adj3 support).mp.	153

29	((early or earlier) adj3 help*).mp.	86

30	(head start or sure start or headstart or surestart).ab,ti.	226

31	("first 1000 days" or "first thousand days" or "1st thousand days" or "1st 1000 days").mp.	2

32	25 or 26 or 27 or 28 or 29 or 30 or 31	29083

33	20 and 24 and 32	120

Social Policy and Practice <202210>

1	(neighbo?rhood care or neighbo?rhood team*).ab,ti.	93

2	(integrat* adj3 health adj3 (care or service* or team* or group or groups or partners* or organi?ation* or consortium* or collaborat*)).ab,ti.	1538

3	Integrated care partnership.ab,ti.	5

4	Integrated care system.ab,ti.	87

5	Place-based partnerships.ab,ti.	16

6	Provider collaboratives.ab,ti.	12

7	healthcare partnership.ab,ti.	2

8	care partnership.ab,ti.	142

9	community partnership.ab,ti.	103

10	((care or healthcare) adj3 integrated).ab,ti.	2236

11	"health and social care partnership".ab,ti.	54

12	"primary and secondary care integration".ab,ti.	2

13	horizontal integration.ab,ti.	6

14	vertical integration.ab,ti.	31

15	shared care.ab,ti.	178

16	(child* or adolescen* or juvenile* or teenage* or youth* or infant* or boy* or girl* or p?ediatric* or young people or young person* or young adult* or youngster* or minor or minors).ab,jn,ti.	133704

17	"improving the health of children and young people".ab,ti.	4

18	early years.mp.	2715

19	((early or prompt or timely or "in time" or expedient or expeditious or anticipat*) adj3 interven*).mp.	4830

20	((early or prompt or timely or "in time" or expedient or expeditious or anticipat*) adj3 support).mp.	782

21	((early or earlier) adj3 help*).mp.	434

22	(head start or sure start or headstart or surestart).ab,ti.	994

23	("first 1000 days" or "first thousand days" or "1st thousand days" or "1st 1000 days").mp.	4

24	1 or 2 or 3 or 4 or 5 or 6 or 7 or 8 or 9 or 10 or 11 or 12 or 13 or 14 or 15	3790

25	16 or 17	133704

26	18 or 19 or 20 or 21 or 22 or 23	8598

27	24 and 25 and 26	40

Date Run:	31/01/2023 21:44:15

Comment:	Cochrane library search history

ID	Search	Hits

#1	MeSH descriptor: [Delivery of Health Care, Integrated] explode all trees	513

#2	MeSH descriptor: [Cooperative Behavior] explode all trees	1098

#3	MeSH descriptor: [Intersectoral Collaboration] explode all trees	66

#4	MeSH descriptor: [Community Health Services] explode all trees	17301

#5	(#2 OR #3) AND #4	185

#6	#1 OR #2 OR #3 OR #5	1647

#7	(neighbo?rhood care OR neighbo?rhood team*):ti OR (neighbo?rhood care OR neighbo?rhood team*):ab	426

#8	(integrat* NEAR/3 health NEAR/3 (care or service* or team* or group or groups or partners* or organi?ation* or consortium* or collaborat*)):ti OR (integrat* NEAR/3 health NEAR/3 (care or service* or team* or group or groups or partners* or organi?ation* or consortium* or collaborat*)):ab	678

#9	("Integrated care partnership" OR "Integrated care system" OR "Place-based partnerships" OR "Provider collaboratives" OR "healthcare partnership" OR "care partnership" OR "community partnership"):ti OR ("Integrated care partnership" OR "Integrated care system" OR "Place-based partnerships" OR "Provider collaboratives" OR "healthcare partnership" OR "care partnership" OR "community partnership"):ab	94

#10	((care or healthcare) NEAR/3 integrated):ti OR ((care or healthcare) NEAR/3 integrated):ab	2153

#11	("health and social care partnership" OR "primary and secondary care integration" OR "horizontal integration" OR "vertical integration" OR "shared care"):ti OR ("health and social care partnership" OR "primary and secondary care integration" OR "horizontal integration" OR "vertical integration" OR "shared care"):ab	252

#12	#6 OR #7 OR #8 OR #9 OR #10 OR #11	4611

#13	MeSH descriptor: [Child] explode all trees	70488

#14	MeSH descriptor: [Adolescent] explode all trees	121283

#15	MeSH descriptor: [Infant] explode all trees	39487

#16	(child* or adolescen* or juvenile* or teenage* or youth* or infant* or boy* or girl* or p?ediatric* or young people or young person* or young adult* or youngster* or minor or minors):ti OR (child* or adolescen* or juvenile* or teenage* or youth* or infant* or boy* or girl* or p?ediatric* or young people or young person* or young adult* or youngster* or minor or minors):ab	243739

#17	#13 OR #14 OR #15 OR #16	338301

#18	MeSH descriptor: [Early Intervention, Educational] explode all trees	606

#19	MeSH descriptor: [Early Medical Intervention] explode all trees	492

#20	("early years" OR "head start" OR "sure start" or headstart OR surestart OR "first 1000 days" OR "first thousand days" OR "1st thousand days" OR "1st 1000 days"):ti OR ("early years" OR "head start" OR "sure start" or headstart OR surestart OR "first 1000 days" OR "first thousand days" OR "1st thousand days" OR "1st 1000 days"):ab	662

#21	((early or prompt or timely or "in time" or expedient or expeditious or anticipat*) NEAR/3 interven*):ti OR ((early or prompt or timely or "in time" or expedient or expeditious or anticipat*) NEAR/3 interven*):ab	8428

#22	((early or prompt or timely or "in time" or expedient or expeditious or anticipat*) NEAR/3 support*):ti OR ((early or prompt or timely or "in time" or expedient or expeditious or anticipat*) NEAR/3 support*):ab	1434

#23	#18 OR #19 OR #20 OR #21 OR #22	10874

#24	#12 AND #17 AND #23	65

*N.B. all papers from Cochrane clinical trials register*

**Cinahl Ultimate search 31-01-2023**

S17	S8 AND S11 AND S16	406

S16	S12 OR S13 OR S14 OR S15	64,052

S15	TI ( (early or prompt or timely or "in time" or expedient or expeditious or anticipat*) N3 support* ) OR AB ( (early or prompt or timely or "in time" or expedient or expeditious or anticipat*) N3 support* )	12,328

S14	TI ( (early or prompt or timely or "in time" or expedient or expeditious or anticipat*) N3 interven* ) OR AB ( (early or prompt or timely or "in time" or expedient or expeditious or anticipat*) N3 interven* )	34,262

S13	TI ( "early years" OR "head start" OR "sure start" or headstart OR surestart OR "first 1000 days" OR "first thousand days" OR "1st thousand days" OR "1st 1000 days" ) OR AB ( "early years" OR "head start" OR "sure start" or headstart OR surestart OR "first 1000 days" OR "first thousand days" OR "1st thousand days" OR "1st 1000 days" )	3,848

S12	(MH "Early Intervention+") OR (MM "Early Childhood Intervention")	20,690

S11	S9 OR S10	1,439,380

S10	TI ( child* or adolescen* or juvenile* or teenage* or youth* or infant* or boy* or girl* or p?ediatric* or young people or young person* or young adult* or youngster* or minor or minors ) OR AB ( child* or adolescen* or juvenile* or teenage* or youth* or infant* or boy* or girl* or p?ediatric* or young people or young person* or young adult* or youngster* or minor or minors )	931,218

S9	(MH "Adolescence+") OR (MH "Child+") OR (MH "Infant+")	1,119,577

S8	S1 OR S2 OR S3 OR S4 OR S5 OR S6 OR S7	51,715

S7	TI ( "health and social care partnership" OR "primary and secondary care integration" OR "horizontal integration" OR "vertical integration" OR "shared care" ) OR AB ( "health and social care partnership" OR "primary and secondary care integration" OR "horizontal integration" OR "vertical integration" OR "shared care" )	1,305

S6	TI ( (care or healthcare) N3 integrated ) OR ( (care or healthcare) N3 integrated )	24,830

S5	TI ( integrat* N3 health N3 (care or service* or team* or group or groups or partners* or organi#ation* or consortium* or collaborat*) ) OR AB ( integrat* N3 health N3 (care or service* or team* or group or groups or partners* or organi#ation* or consortium* or collaborat*) ) OR AB ( "Integrated care partnership" OR "Integrated care system" OR "Place-based partnerships" OR "Provider collaboratives" OR "healthcare partnership" OR "care partnership" OR "community partnership" ) OR TI ( "Integrated care partnership" OR "Integrated care system" OR "Place-based partnerships" OR "Provider collaboratives" OR "healthcare partnership" OR "care partnership" OR "community partnership" )	8,557

S4	TX (neighbo#rhood W1care) or (neighbo#rhood W1 team*)	204

S3	((MH "Community Health Services+") OR (MH "Community Mental Health Services+")) AND ((MM "Cooperative Behavior") OR (MM "Collaboration"))	3,312

S2	(MM "Cooperative Behavior") OR (MM "Collaboration")	22,512

S1	(MM "Health Care Delivery, Integrated")	9,659

**ProQuest Psyc Info search: 31/01/2023**

((MAINSUBJECT.EXACT.EXPLODE("Integrated Services") OR (MAINSUBJECT.EXACT.EXPLODE("Cooperation") OR MAINSUBJECT.EXACT.EXPLODE("Cotherapy"))) OR MAINSUBJECT.EXACT.EXPLODE("Collaboration") OR (MAINSUBJECT.EXACT.EXPLODE("Community Mental Health Services") AND MAINSUBJECT.EXACT.EXPLODE("Integrated Services")) OR (abstract(neighbo?rhood care OR neighbo?rhood team*) OR title(neighbo?rhood care OR neighbo?rhood team*)) OR (abstract(integrat* NEAR/3 health NEAR/3 (care OR service* OR team* OR group OR groups OR partners* OR organi?ation* OR consortium* OR collaborat*)) OR title(integrat* NEAR/3 health NEAR/3 (care OR service* OR team* OR group OR groups OR partners* OR organi?ation* OR consortium* OR collaborat*))) OR (tiab("Integrated care partnership" OR "Integrated care system" OR "Place-based partnerships" OR "Provider collaboratives" OR "healthcare partnership" OR "care partnership" OR "community partnership") OR tiab((care OR healthcare) NEAR/3 integrated) OR tiab("health and social care partnership" OR "primary and secondary care integration" OR "horizontal integration" OR "vertical integration" OR "shared care"))) AND (child* OR adolescen* OR juvenile* OR teenage* OR youth* OR infant* OR boy* OR girl* OR p?ediatric* OR young people OR young person* OR young adult* OR youngster* OR minor OR minors) AND (MAINSUBJECT.EXACT.EXPLODE("Early Intervention") OR tiab("early years" OR "head start" OR "sure start" OR headstart OR surestart OR "first 1000 days" OR "first thousand days" OR "1st thousand days" OR "1st 1000 days") OR tiab((early OR prompt OR timely OR "in time" OR expedient OR expeditious OR anticipat*) NEAR/3 interven*) OR tiab((early OR prompt OR timely OR "in time" OR expedient OR expeditious OR anticipat*) NEAR/3 support*))

**475 hits total**
